# Coronary Artery Disease (CAD) - risk factors and causes: In Libyan patients

**DOI:** 10.1101/2021.11.18.21266513

**Authors:** Osama Bheleel, Alaa Abdulhamid, Ibtisam Alhadi, Hanaa Grash, Hajer almuaket, Mohamed Hadi Mohamed Abdelhamid

**Author notes:** **Corresponding author:** Mohamed Hadi Mohamed Abdelhamid, National Center of Disease Control (NCDC); Department of Cell Biology and Tissue Culture, Biotechnology Research Center (BTRC), Tripoli - Libya; Phone N: 00 218 916030276. **Funding** This research did not receive any specific grant from funding agencies in the public, commercial, or not-for-profit sectors.

## Abstract

**Aims:** Coronary artery disease (CAD) is the leading cause of death worldwide in both men and women. Accordingly, we retrospectively reviewed the effects of various risk factors on coronary angiographic outcomes.

**Methods and Results:** Data were collected from the catheter lab through Tripoli university hospital records, whereas the team reviewed clinical data and coronary artery diagrams for one year from 01/04/2019 to 31/03/2020. In our study, the total number of cases was 666; 401 male and 265 female, ranging in age between 27 and 91 years. Considering the data, a significantly increased incidence of coronary artery disease (CAD) among the male who smokes, and who were less than 60 years of age. Furthermore, in the present study, the total number of normal patients was predominantly female. The most common risk factors for women were diabetes mellitus (DM), hypertension (HTN) (12%,13% respectively). While the men share the significant effects of smoking on coronary angiography (C. Angio) findings (40.52%), and most of them underwent a percutaneous coronary intervention (PCI).

**Conclusions:** In our study, there was evidence that CAD is a prevalent disease among the middle-aged populations with male gender preference. The risk factors including diabetes, hypertension, and smoking are the most contributing factors for the developing CAD in Libya.

## Introduction

Coronary artery disease (CAD) is the leading cause of death worldwide in both men and women. Approximately 1 in 30 patients with stable CAD experiences cardiovascular death or myocardial infarction (MI) each year ^1^. It is the third leading cause of mortality worldwide and is associated with 17.8 million deaths annually. As well it places a large economic burden on the community ^1–3^. Moreover, there is little literature related to the epidemiology of CAD in Africa. Though, changes in lifestyle and modernization have risen the prevalence of CAD. Previous studies reported that the incidence of coronary artery disease (CAD) is rising by 160% in the Middle East and North Africa, and the mortality rate from CAD was high (120 per 100,000 populations) ^4,5^. As in Tunisia, where studies have reported coronary artery disease (CAD) mortality rates increased by 11.8% for men and 23.8% for women, between 1997 and 2009, further causing the death of 70% of cardiovascular patients ^6,7^. In addition, coronary artery disease has reached epidemic proportions in Egypt, the mortality rates due to CAD was measured to be 280 per 100,000 populations^8^.

There are several risk factors for CAD, some can be controlled but not all. The non-modifiable risk factors are as follows; age, sex, race, family history, and/or physio-pathological conditions ^9,10^.

Moreover, according to the 52 - country case-control study, nine easily measurable, controllable, and/or modifiable risk factors account for more than 90% of the heart diseases risk ^11^. These nine risk factors include smoking, abnormal blood lipid levels, high blood pressure, diabetes, obesity, a lack of physical activity, junk food, alcohol overconsumption, and the psychosocial index ^11,12^. Among these cases, the most common form of cardiovascular disease, with an estimated prevalence of CAD in men than women ^11^. In additionally, several studies showed that modifiable classical risk factors for CAD, except for smoking, were more prevalent in women and were associated with their diet ^13,14^.

In fact, there are three main strategies are available for angina control and prevention or reversal of plaque progression: medical treatment, Percutaneous coronary intervention PCI, and/ or Coronary artery bypass grafting CABG^15^.

Percutaneous coronary intervention (PCI) is often part of standard therapy in patients presenting with significant coronary artery disease (CAD). In the same vein, in patients with stable CAD, PCI can be considered a valuable initial mode of revascularisation in all patients with objective large ischaemia in the presence of almost every lesion subset, with only one exception: chronic total occlusions that cannot be crossed ^16^.

Since 1977, when Grundzig performed the first PTCA in Zurich, percutaneous coronary intervention has been recognized as a leading procedure in treating coronary artery disease patients and has become a common part of routine practice worldwide ^17^. Correspondingly, Coronary artery bypass grafting (CABG) is an acceptable procedure used to treat coronary artery disease, also management of refractory to medical treatment. Globally, there are around one million patients to undergo coronary artery bypass graft (CABG) surgery each year ^18^.

In Libya, there is rapid expansion and uptake of coronary angiography (C. Angio) procedure. However, when taking the entire clinical practice in Libya into consideration, limited data are available to describe nationwide contemporary practice patterns of C. Angio. Yet, until now, no unified nationwide registry has been established to demonstrate the accurate figures of cardiac procedures; Despite this, there has been a limited routine collection of related data particularly around quality, safety, and cost. With respect to the growing need for C. Angio, the development of C. Angio registries in Libya was of growing interest. As a clinical quality registry, a C. Angio database is an important mechanism of monitoring and benchmarking the performance of clinical care, improving safety and outcomes, contributing to reducing treatment costs, and regulating guidelines.

In this context, the objective of this study was to describe the characteristics and outcome of consecutive unselected Libyan patients who underwent diagnostic C. Angio in a nationwide cohort of one year. The present study is the first part of a study of coronary artery disease risk factors and causes in Libyan patients.

## Material and Method

### Study population

The present study is the first part of a study of coronary artery disease risk factors and causes in Libyan patients. The total number of patients records reviewed who underwent diagnostic C. Angio during this study was 666 cases, however, 51 cases were dropped due to personal reasons hence the data of these cases were excluded. Therefore, 615 cases were contributed to this study: 371 male and 244 female. The rang age of the study population was 27-91 years. Coronary Angio patients’ reports were revised in the Cath Lab of Tripoli University Hospital.

### Data collection

The data was collected from the catheter lab in Tripoli University Hospital records, the team reviewed all clinical data of patients such as; (BMI, history family, lipid profiles, age, gender, chronic diseases, and others), we also reviewed all coronary artery diagrams during one year from 01/04/2019 to 31/03/2020 retrospectively. The axis to the files of the patients and their contact details was extremely difficult. Furthermore, there are clinical data is missing such as physical activity, the behavior of feeding, and physio-pathological conditions.

### Ethics statement

The study was approved by the Ethics Committee (Libyan National committee for Biosafety and Bioethics) and the protocol has been previously ^19^. An informed consent was obtained in all patients; and the study was carried out according to Helsinki declaration.

### Statistical analysis

Continuous variables were presented as mean ± SD and were compared using Two-way analysis of variance. We divided the cases age to three group (age; ≤40, 40-60 and ≥60 yeas) and two generals risk factors (modifiable, non-modifiable risk factor). In all cases, p values <0.05 were considered statistically significant. All data were processed using the Statistical Package for Social Sciences, version 25 (SPSS, Chicago, IL, USA)

## Results

The total number of cases is 615; 371 (60.32 %) men and 244 (39.67 %) women, The mean age of the study population was 59.4 ± 13.4 years (an age range, 27–91 years). We have recorded that no classical risk factors with 276 patients, representing 44% of total cases. Furthermore, these cases were dilated cardiomyopathy, left bundle branch block, and preoperative assessment for cardiac surgery.

### Non-modifiable risk factors for CAD

The findings of this study showed that the percentage of men who underwent C. Angio was (55.19 ± 10.2; P ≤ 0.05). There was Also, a significant difference was found between the two genders with respect to the number of underwent C. Angio in favor of men. The results indicate that ages were mostly between 40 and 60 years old (52.92%). At this age group, the percentage of cases was significantly higher for men compared to women (56.42% versus 48.55%; P ≤ 0.05). The demographic characteristics and clinical data of the participants are summarized in Table 1.

**Table 1.** Prevalence of risk factors stratified by modifiable and Non-modifiable risk factors. N: Number of patients who have risk factors; HTN: hypertension; DM: diabetes mellitus; (*): High significant different (a p-value <0.05 was considered significant); (**): Very high significant different (a p-value <0.05 was considered significant).

### Modifiable risk factors for CAD

According to the results of this study, the most common risk factors for women were diabetes mellitus (DM), hypertension (HTN), and women who have DM + HTN (12.8%,13.9%, 52.17% respectively), however no cases of smoke have been reported among women (Table. 1).

The prevalence intensity of men risk factors was as follows: smoking (36.7%), HTN (14.11%) and DM (12.04 %), which were lower compared to women’s cases at the last two factors (Table. 1). The incidence of smokers was significantly higher than the other risk factors.

### Result of coronary angiography (C. Angio)

The results of C Angio showed that the number of cases that do not require any medical or surgical intervention was 186 (30%), of whom the largest percentage of them were women. While the data showed that the percentage of men who underwent Medical, PCI, and CABG were (24.74%, 40.97%, and 14.55% respectively). The incidence of PCI cases was one and a half-time for men than for women (Fig.1). The procedure of PCI and CABG was increasing with age and includes age >40 years in men and >60 years in women (Fig.2)

**Fig 1.**
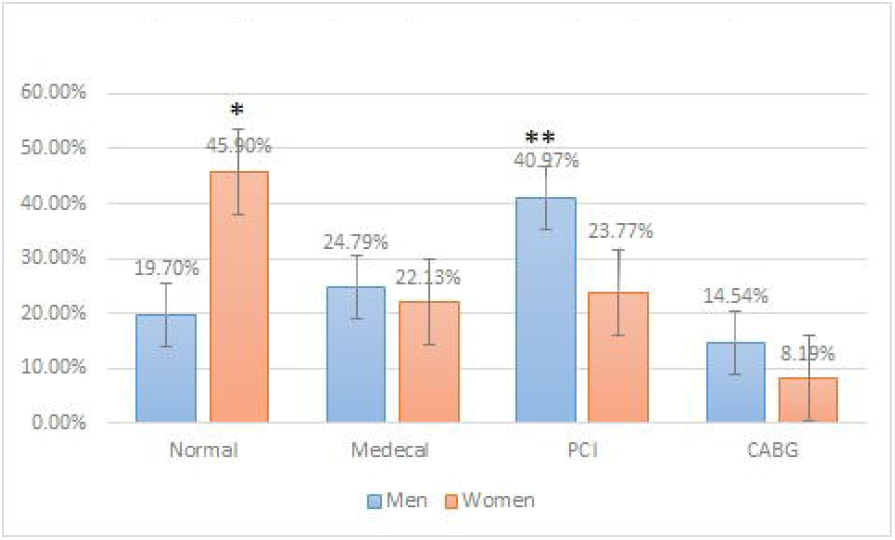
Effect of gender on resulte of C. Angio. PCI: percutaneous coronary intervention; CABG: Coronary artery bypass grafting; (*): High significant different (a p-value <0.05 was considered significant); (**): Very high significant different (a p-value <0.05 was considered significant).

**Fig 2.**
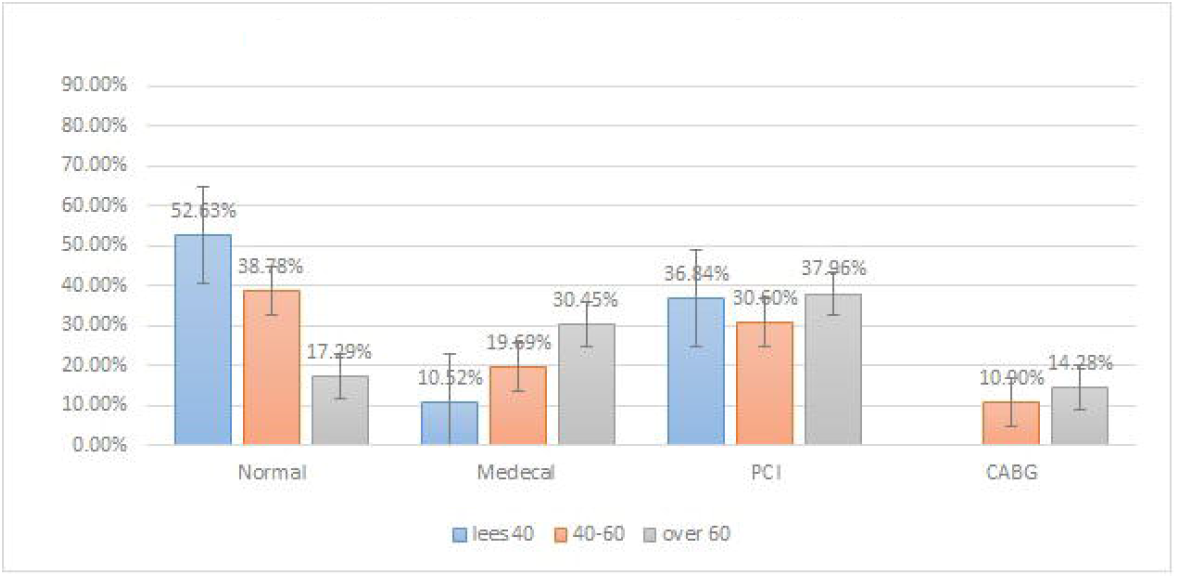
Effect of age factor on result of C. Angio. PCI: percutaneous coronary intervention; CABG: Coronary artery bypass grafting.

Furthermore, our data show that Percutaneous coronary intervention (PCI) was significantly influenced by risk factors than the others. Approximately 15% of all Coronary artery bypass grafting (CABG) were performed on men with diabetes. While strong evidence was found that men who underwent a PCI were smokers (42%) (a p-value <0.05 was considered significant) (Fig. 3).

**Fig 3.**
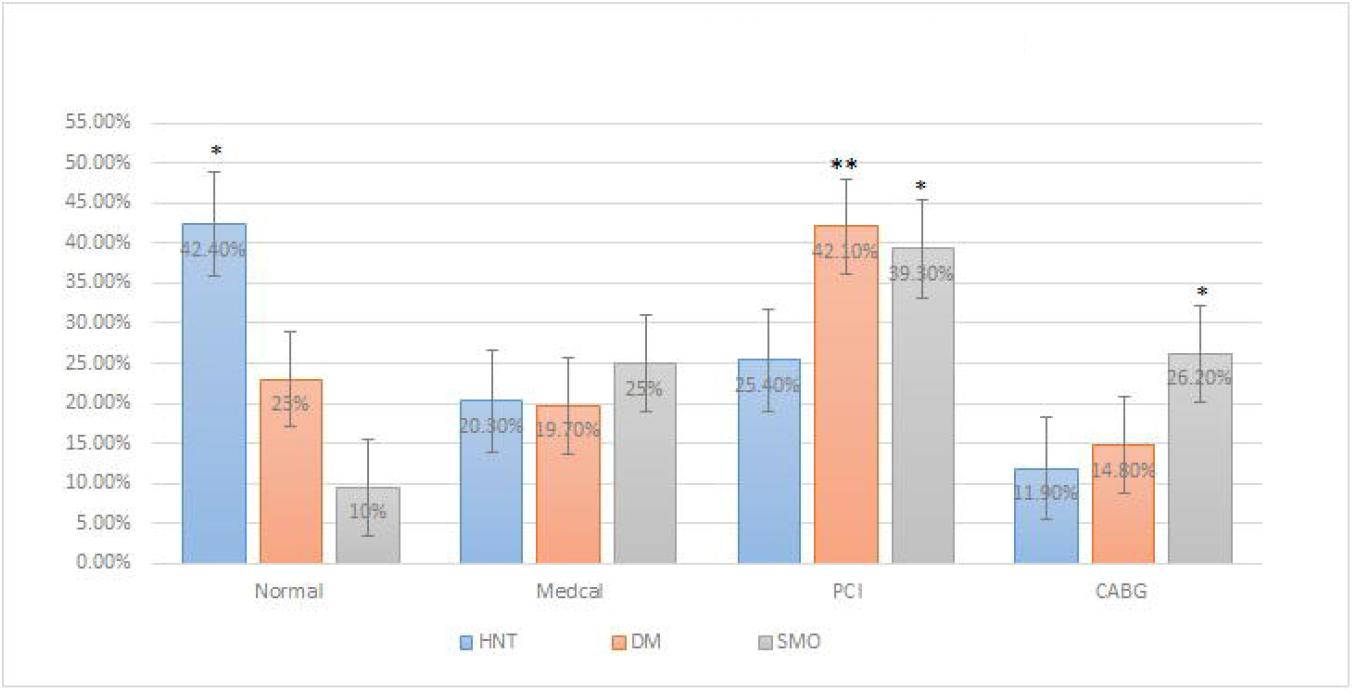
C. Angio results with different risk factors. PCI: percutaneous coronary intervention; CABG: Coronary artery bypass grafting; (*): High significant different (a p-value <0.05 was considered significant); (**): Very high significant different (a p-value <0.05 was considered significant).

## Discussion

The current study represents the first Libyan enrollment involving CAD and the result of C. Angio for patients managed at the public sector, in university and regional hospitals. Over half of the patients who underwent C. Angio were men (60.32%), which is to be expected because CAD is a primarily men gender disease. This is consistent with the data obtained by Adda et al (2019), who found that patients who underwent C. Angio are more men than women (88.5% vs 21.5% respectively) ^6^.

The second strong observation is that most of the cases that underwent of C. Angio fall into the age group between 40 and 60 years, and after that the patients with age over 60 years. In the same vein, the hypothesis by different previous studies showed that in women under the age of 55, the incidence of CAD is one-third that of men. However, it gradually increases with age when crossing over 55 years. This hypothesis has been generally accepted^20–22^.

Since the physiological differences between two genders suggest that age-related changes in endogenous sex hormones, such as estrogen, progesterone, and androgens, play a pivotal role in controlling CAD. Exceptionally, the hormone estrogen plays essential acting as a cofactor in the onset of CAD in women ^23,24^.

The CAD is the disease of the middle age group which is the product group in the community. This fact has an impact on the productivity and performance of the age group mentioned earlier, in the matter of fact, this will be reflected on the community in various aspects as absences from work, the cost of the health care services, expensive medical and device therapy, and the future outcome in the form of chronic heart failure and its sequel. Those facts highlight the prevention strategies and education of the community starting at early age groups especially among adolescents aiming to reduce the burden of the CAD is the cornerstone in any health strategies for the ministry of health.

Moreover, CAD was affecting all groups of the study as a result of modernization associated with the modifiable risk factors for CVD, including diabetes, high blood pressure, smoke, and both risk factors.

In addition, more than half of our population sample had at least one major risk factor. Regarding the risk factors for CAD, about one-third of the cases were diabetics and hypertension or were having both. As both diabetes mellitus and hypertension are prevalent diseases in our community, the tighter glycemic control of diabetics and targeting the high blood pressure will result in a reduction of the incidence and prevalence of CAD in the community ^25–27^.

So far, in men, smoking is one of the major risk factors for cardiovascular diseases causes and is a leading cause of death. This effect is very prevalent. It should be noted that the percentage of Libyan smokers aged between 25 – 64 years among men was very high at 49.6% ^28^. We believe that the high level of Libyan smokers resulted in an increase in the number of men who underwent C. Angio in our study. It also appears that the exposure to multi-factors at the same time increases the requiring medical or PCI intervention if compared to normal C. Angio result.

Considering the results of the C. Angio in our studied population, over 40% of the studies revealed normal coronary angiography, most of them were women (women patients who underwent the C. Angio have normal epicardial coronary arteries), this again highlights that screening for CAD among female gender ought to be reevaluated with the aim to use more non-invasive modalities of screening for CAD such as stress echocardiography or CT-coronary angiography and not to depend only on exercise ECG for sending patients for C. Angio as it has high false-positive results. A 34.1% of the cases underwent PCI implantation, the most stented single artery is left anterior descending artery 12.8% (data not shown).

### Limitations of study

Since our study is a retrospective study, there were difficulties in obtaining the data of the patients. There is no archiving system for the results of the coronary angiograms, and the personal data of the patients were not properly taken. The contact telephone numbers were missing. Most of the patient’s data regarding the cardiac risk factors are not recorded. Furthermore, the details of the procedure as the approach either femoral or radial, the medications during the procedure, the complications either immediate or remote are not registered. In the place of the study, no electronic archiving of the patient information is available, depending only on paper archiving which was not organised professionally, all those facts have a negative impact on our research team.

## Conclusion

In our study, there was evidence that CAD is a prevalent disease among the middle-aged populations with male gender preference. Clinical data indicated that the Libyan adult population is of a high level of CAD risk factors including diabetes, hypertension, and smoking are the most contributing factors for the development of CAD, those risk factors are modifiable and preventable direct causes of CAD, which may require urgent decision-making to address national control measures regarding CAD. The national registry of the coronary procedure is crucial and mandatory for patient safety and to have quality control of the services provided in the cath labs.

## Data Availability

All data produced in the present work are contained in the manuscript

## Acknowledgements

The authors wish to thank all doctors, nurses, and technicians in the Department of Cardiology at Tripoli University Hospital who were involved in this study, and Hamza El-thelb for editing the text.

## Contributors

(I)Manuscript writing: All authors; (II) Final approval of manuscript: All authors.

